# The Role of Diet on the Gut Microbiome, Mood and Happiness

**DOI:** 10.1101/2023.03.18.23287442

**Authors:** Sydney E. Martin, Colleen S. Kraft, Thomas R. Ziegler, Erin C. Millson, Lavanya Rishishwar, Greg S. Martin

## Abstract

The gut microbiome may be both helpful and harmful, and not only is it affected by diet, it has also been shown to affect mental health including personality, mood, anxiety and depression. In this clinical study we assessed dietary nutrient composition, mood, happiness, and the gut microbiome in order to understand the role of diet in the gut microbiome and how that affects mood and happiness. For this pilot study, we enrolled 20 adults to follow this protocol: recording a 2-day food log, sampling their gut microbiome, and completing five validated surveys of mental health, mood, happiness and well-being, followed by a minimum 1 week diet change and repeating the food log, microbiome sampling and the 5 surveys. The change from a predominantly Western diet to vegetarian, Mediterranean and ketogenic diets led to changes in calorie and fiber intake. After the diet change, we observed significant changes in measures of anxiety, well-being and happiness, and without changes in gut microbiome diversity. We found strong correlations between greater consumption of fat and protein to lower anxiety and depression, while consuming higher percentages of carbohydrates was associated with increased stress, anxiety, and depression. We also found strong negative correlations between total calories and total fiber intake with gut microbiome diversity without correlations to measures of mental health, mood or happiness. We have shown that changing diet affects mood and happiness, that greater fat and carbohydrate intake is directly associated with anxiety and depression and inversely correlated with gut microbiome diversity. This study is an important step towards understanding how our diet affects the gut microbiome and in turn our mood, happiness and mental health.

## Introduction and Background

Microbes begin living in the gut shortly after birth and are vital to a healthy immune system. Initial development of the microbiome comes from the placenta, amniotic fluid, meconium and vaginal exposure during birth. After birth, breastfeeding increases *Bifidobacterium*, which increases immunoglobulin A (IgA) and contributes to gut immunity, and decreases levels of the pro-inflammatory cytokine IL-6 which manifests with both acute and chronic inflammation. Healthy gut function has been linked to normal central nervous system function.^1^

The adult microbiome is made up of primarily *Bacteroidetes* and *Firmicutes* with smaller numbers of *Proteobacteria, Verrucomicrobia, Actinobacteria*, and *Cyanobacteria phyla*, and *Fusobacteria genus*. Irrespective of variability between individuals, there are three enterotypes based on a person’s dominant bacterial composition: *Bacteroides, Prevotella*, or *Ruminococcus*. The dominant species are determined by dietary intake, such that the *Prevotella* enterotype is associated with diets high in carbohydrates (e.g. Mediterranean diet), *Bacteroides* is predominant in people eating large amounts of protein (e.g. Western diet) and *Ruminococcus* is the most common enterotype overall and is seen in people eating a mixture of proteins and simple sugars. These enterotypes are independent of environmental factors like age, body-mass index, gender, and geographic location and are primarily determined by diet and genetics.

Many factors play a role in producing the gut microbiome including diet, environment, season, and health status. When the human microbiome is challenged with changes in diet, stress, or antibiotics, the physiology of the microbiome changes. This change may lead to increased gut permeability and allows contents such as bacterial metabolites (e.g. endotoxin or lipopolysaccharide, LPS) and bacteria themselves to move from the gut lumen into the circulation, called “leaky gut syndrome.” The abnormal gut permeability promotes local mucosal and systemic inflammation, which results in the release of cytokines and neurotransmitters that in turn may worsen gut permeability, creating a vicious cycle. These chemical mediators can influence brain function via the gut-brain axis (GBA) and lead to anxiety, depression and memory loss.

The gut microbiota can regulate emotions through the gut-brain axis (GBA).^2,3^ The GBA was initially discovered by Sudo in germ-free mice, and since then studies have shown that the GBA extends into the endocrine, neural, and immune pathways.^4,5^ Human studies have shown an increase in gut bacterial translocation in mood disorders.^6,7^ For example, in major depressive disorder there are significant increases in *Bacteroidetes, Proteobacteria*, and *Actinobacteria* and decreases in *Firmicutes* compared to controls. There is a strong association between bacterial group and neurocognitive reactions to stressful images, and increased *Bacteroides* enterotype and increased gray matter in the frontal lobes, cerebellum and hippocampus whereas *Prevotella* predominance is associated with greater white matter in attentional, emotional, and sensory processing areas. Ninety percent (90%) of serotonin receptors are located in the gut, and the most common pharmacologic treatment for mood disorders, selective serotonin reuptake inhibitors or SSRI’s has primarily gastrointestinal side effects. There is anatomical and physiologic two-way communication between the gut and brain via the vagus nerve, including enteroendocrine cells recently discovered in the gut.^8^ The GBA explains many connections between diet and psychological diseases such as depression and anxiety, but also has led to the creation of the new field of nutritional psychiatry to therapeutically treat patients by nutritional alterations.^9-11^

The composition and amount of nutritional intake influences mood and happiness. Consumption of protein slows absorption of carbohydrates and increases the release of dopamine and norepinephrine, which has direct effects on mood. Similarly, eating carbohydrates increases serotonin, which also has direct effects on mood. In addition, the chemical acetylcholine is more present in wheat germ and eggs and is directly involves in neurotransmission and has been associated with learning, memory and mood. Omega-3 fatty acids affect mood and behavior, and low blood levels are associated with depression and pessimism.^12^ Certain foods have been described in the lay press as “brain super foods” or affecting cognition and mood: Brazil nuts, oily fish that are high in omega-3 fatty acids, oats, bananas, lentils, chicken and turkey, spinach, quinoa, and dark chocolate.^13^ It has also been shown that changes in mood can alter food preferences, with people who are feeling sad more often choosing “comfort foods” rather than healthy alternatives.^14^

Changes in diet may affect mood and happiness through the gut microbiome. Changes in diet have been shown to account for 57% of gut microbiome variation in mice, while genetic background contributed only 12%.^15^ In the Nature journal series, the ketogenic diet was found to produce changes in taxonomic and functional composition of the microbiome in children with epilepsy.^16^ A similar study in childhood epilepsy found changes in the proportional representation of gut microbiome enterotypes.^17^ The benefits of ketogenic diets for controlling severe epilepsy may be mediated by the gut microbiome, as the effects can be reproduced in animal models by direct manipulation of gut bacterial enterotypes without changing diet.^18^ Similarly, the Mediterranean diet causes a higher ratio of *Bacteroidetes* to *Firmicutes*, and high consumption of animal protein, saturated fats, and sugars affect gut microbiota diversity.^19-23^ Changing diet has been shown to alter the gut microbiome within 24 hours.^24^

We aimed to determine in a pilot study the association of gut microbiome diversity and proportional enterotype representation with dietary nutrient composition and with measures of mood and happiness. We hypothesized that the diversity of organisms in the gut microbiome and the distribution of each major microbiome organism enterotype will associate with diet composition and with measures of mood and happiness.

## Materials and Methods

This study protocol was approved by the Emory University Institutional Review Board and the Georgia Clinical and Translational Science Alliance Clinical Research Centers (Georgia CTSA GCRC) Scientific Advisory Committee. An application was submitted to the uBiome company who agreed to provide microbiome testing kits and data analysis at a reduced price. For each study subject, eligibility was confirmed and informed consent was obtained per policy of the Emory University IRB and the U.S. Code of Federal Regulations.

### Subject eligibility

#### Inclusion criteria

adults > 18 years of age maintaining a consistent and unchanging diet for at least 3 days prior to the time of participation

#### Exclusion criteria

acute illness, antibiotic therapy in the past 90 days, known or suspected gastrointestinal disorder, clinical depression or similar disorders (e.g. dysthymia or bipolar affective disorder), weight gain or loss of more than 5% of body weight in the past 90 days, pregnancy, vulnerable individual (cognitive impairment, ongoing mental or psychiatric illness, prisoner).

### Study Procedures

The study consisted of each subject completing surveys, biometric and nutritional information described below as part of study data collection, and they were provided one uBiome gut microbiome testing kit with information on how to collect a sample at home to submit in a pre-paid and pre-addressed mailing kit from uBiome (**Figure 1**). After completing the baseline assessment, subjects who wished to alter their previous diet were given an addition uBiome gut microbiome testing kit and instructions to repeat the same assessments (surveys, biometric and nutritional information) a minimum of 5 days after a dietary change of their choosing.

**Figure 1.**
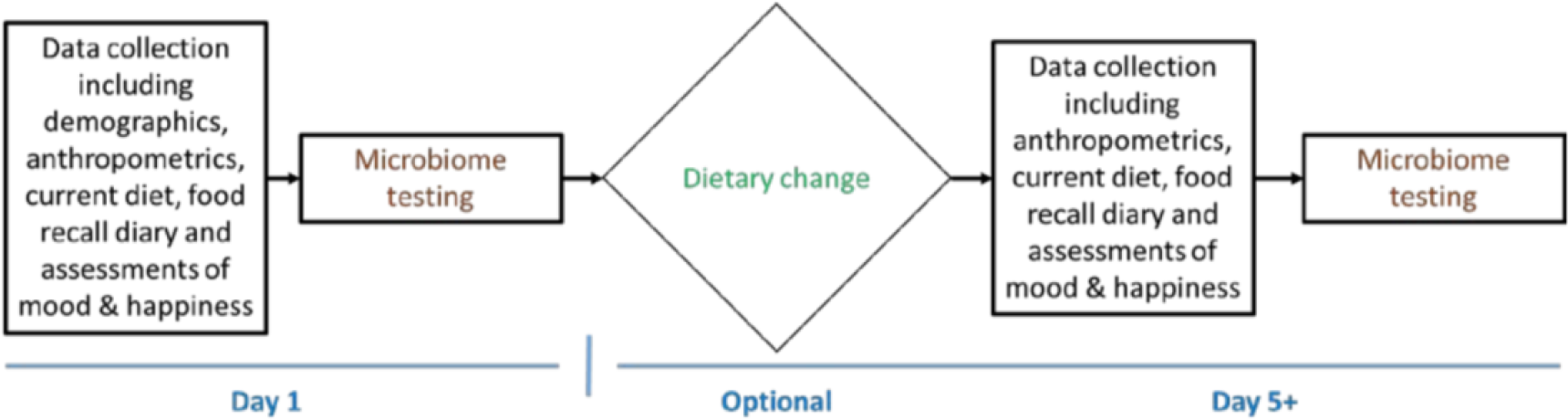
Timeline for participant completion of the study.

### Data collection

We recorded general self-reported demographics and anthropometrics such as age, gender, race, ethnicity, height, and weight. We also recorded general health assessments with a review of medical diagnoses and medical symptoms, particularly focusing on the gastrointestinal and psychological systems described in the eligibility criteria. For the dietary assessment, each participant created a two-day food record that was analyzed by a registered dietician using the University of Minnesota Nutrition Data System for Research (NDSR) for determining core nutritional components. For this study, the NDSR nutrient extraction focused on total calories, proportions of fat, carbohydrates, and protein, and the total fiber intake during the two-day food record. Psychological surveys and information about current diet were assessed using a secure website for electronic data capture. The five validated mood surveys were the Affectometer-2^25^; Warwick Edinburgh Mental Well-Being Scale (WEMWBS)^26^; WHO-5 Well-Being Index^27^; Depression, Anxiety, and Stress Scale (DASS-42)^28^; and Hospital Anxiety and Depression Scale (HADS)^29^. The five mood surveys measured the happiness, satisfaction, stress, anxiety, and depression of each participant. The Affectometer-2 measured both positive and negative characteristics of each participant on an overall scale from -80 to +80; the HADS measured on a scale from 0-21 for both anxiety and depression; the DASS-42 measured stress, anxiety, and depression on a scale of 0-42 for each characteristic; the WEMWBS measured happiness and a lack of stress on a scale of 14-70; finally, the WHO-5 measured overall satisfaction on a scale from 0.00-1.00.

### Statistical analysis

We first conducted descriptive analyses of the enrolled subjects, followed by comparisons of nutrition, mood and microbiome diversity from before and after the changes in diet (Chi square and paired t-tests). In order to explore the associations between diet, mood or happiness and the microbiome, we correlated measures of gut microbiome diversity (diversity indices) and diet composition, and with the continuous measures of happiness, mood, anxiety, stress and resilience (Affectometer-2, WEMWBS, WHO-5, DASS-42 and HADS) using Pearson correlation coefficient (r). Because this was a pilot study, we considered significant any p-values less than 0.20 in order to maximize the power of the study to detect potential effects and relationships that warrant further investigation.^30,31^

## Results

This study enrolled 20 adults with a mean age of 48.4 years and 12/20 (60%) of subjects being female. The demographic and anthropometric characteristics of the study subjects are shown in **Table 1**. Following consent, each study participant was given a form for recording their diet for two consecutive days, instructions on how to complete the mood and happiness surveys online, and two uBiome kits with sample collection and kit mailing instructions. The investigators worked with study participants when they had questions or did not complete parts of the study as planned.

**Table 1.**
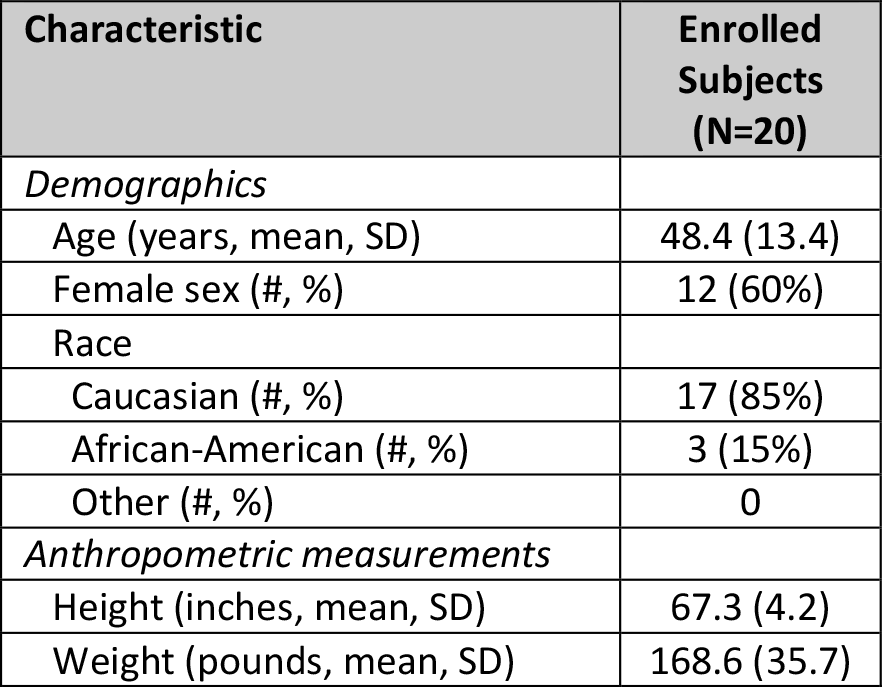
Demographic and Anthropometric Characteristics of Study Subjects.

Most of the participants began the study on a typical Western diet without prebiotics or probiotics, as shown in **Figure 2** and in **Table 2**. During the study, the most frequent diet changes were to a Vegetarian, Mediterranean, or Ketogenic diet, with the highest percentage of participants choosing Vegetarian. Other diets included Weight Watchers and unnamed diets. Secondary analysis created an overview of the changes in diet and mood across the two time points of the diet change. For study subjects as a group, the diet change resulted in a mean reduction in total calories by 218 kcal/day and a reduction in total fiber intake by 4 grams (**Table 2**). The proportions of fat, carbohydrate, and protein intakes remained statistically similar from baseline to follow-up. The number of total calories consumed per day was greater for men than for women both at baseline (2229 vs. 1588 kcal/day, p=0.004) and at follow-up (1929 vs. 1437 kcal/day, p=0.105).

**Table 2.** Nutrition Characteristics of Study Subjects.

| Characteristic | Baseline | Follow-Up | p-value |
| --- | --- | --- | --- |
| *Diet Type* |  |  | 0.73 |
| Western (#, %) | 19 (95%) | 0 |  |
| Vegetarian (#, %) | 0 | 8 (40%) |  |
| Mediterranean (#, %) | 0 | 3 (15%) |  |
| Ketogenic (#, %) | 1 (5%) | 4 (20%) |  |
| Other (#, %) | 0 | 5 (25%) |  |
| Prebiotic/Probiotic use (#, %) | 1 (5%) | 2 (10%) | 0.548 |
| Total calories (mean kcal, SD) | 1844 (470) | 1626 (529) | 0.099 |
| Caloric breakdown |  |  |  |
| Proportion fat intake (% ,SD) | 40.8% (6.8) | 37.5% (10.8) | 0.316 |
| Proportion carbohydrate intake (% ,SD) | 38.8% (8.8) | 37.6% (15.1) | 0.266 |
| Proportion protein intake (% ,SD) | 18.8% (5.1) | 19.2% (8.7) | 0.889 |
| Total fiber intake (mean grams, SD) | 18.9 (9.0) | 14.9 (6.9) | 0.058 |

**Figure 2.**
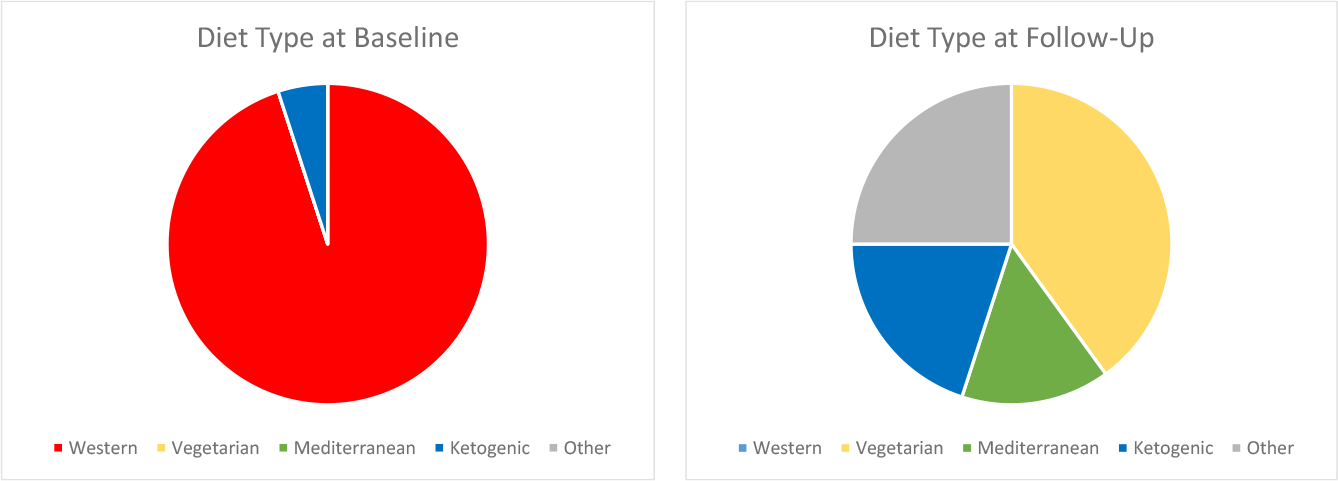
Diet Types at Baseline and at Follow-Up.

Study subjects had typical baseline measures on the mood and happiness surveys, as shown in **Table 3**. From baseline to follow-up we observed consistent changes in the same direction for each mood survey over the duration of the study, including significant p-values for all surveys except for the HADS Depression score. Overall, the average Affectometer-2 score increased by 35% or 11.7 points, WEMWBS increased by 6.7% or 3.5 points, and WHO-5 increased by 11% or 0.07. The HADS Anxiety score decreased by 15% or 1.1 points while HADS Depression remained constant. The DASS-42 Stress score decreased by 27.1% or 3.2 while DASS-42 Anxiety decreased by 46% or 1.6 points and DASS-42 Depression decreased by 41% or 2.0 points (**Table 3, Figure 3**).

**Table 3.** Mental Health, Mood, Happiness and Well-Being Characteristics of Enrolled Subjects.

| Survey instrument | Baseline | Follow-Up | p-value |
| --- | --- | --- | --- |
| *Affectometer-2* |  |  |  |
| Total score (mean, SD) | 33.0 (19.0) | 44.7 (22.0) | 0.014 |
| Positive score (mean, SD) | 54.9 (9.4) | 59.3 (13.4) | 0.113 |
| Negative score (mean, SD) | 21.9 (9.7) | 14.6 (10.0) | 0.003 |
| *Hospital Anxiety and Depression Scale (HADS)* |  |  |  |
| Anxiety (A) score (mean, SD) | 7.5 (2.6) | 6.4 (2.4) | 0.098 |
| Depression (D) score (mean, SD) | 13.8 (1.8) | 13.8 (2.7) | 0.999 |
| *Depression, Anxiety and Stress Scale (DASS-42)* |  |  |  |
| Stress score (mean, SD) | 11.8 (6.7) | 8.6 (7.3) | 0.115 |
| Anxiety score (mean, SD) | 3.5 (4.2) | 1.9 (3.4) | 0.023 |
| Depression score (mean, SD) | 4.9 (5.4) | 2.9 (6.9) | 0.163 |
| *Warwick-Edinburgh Mental Wellbeing Scale (WEMWBS)* |  |  |  |
| Total score (mean, SD) | 52.1 (8.3) | 55.6 (10.0) | 0.137 |
| *WHO-5 Well-Being Index* |  |  |  |
| Total score (mean, SD) | 0.61 (0.17) | 0.68 (0.20) | 0.089 |

**Figure 3.**
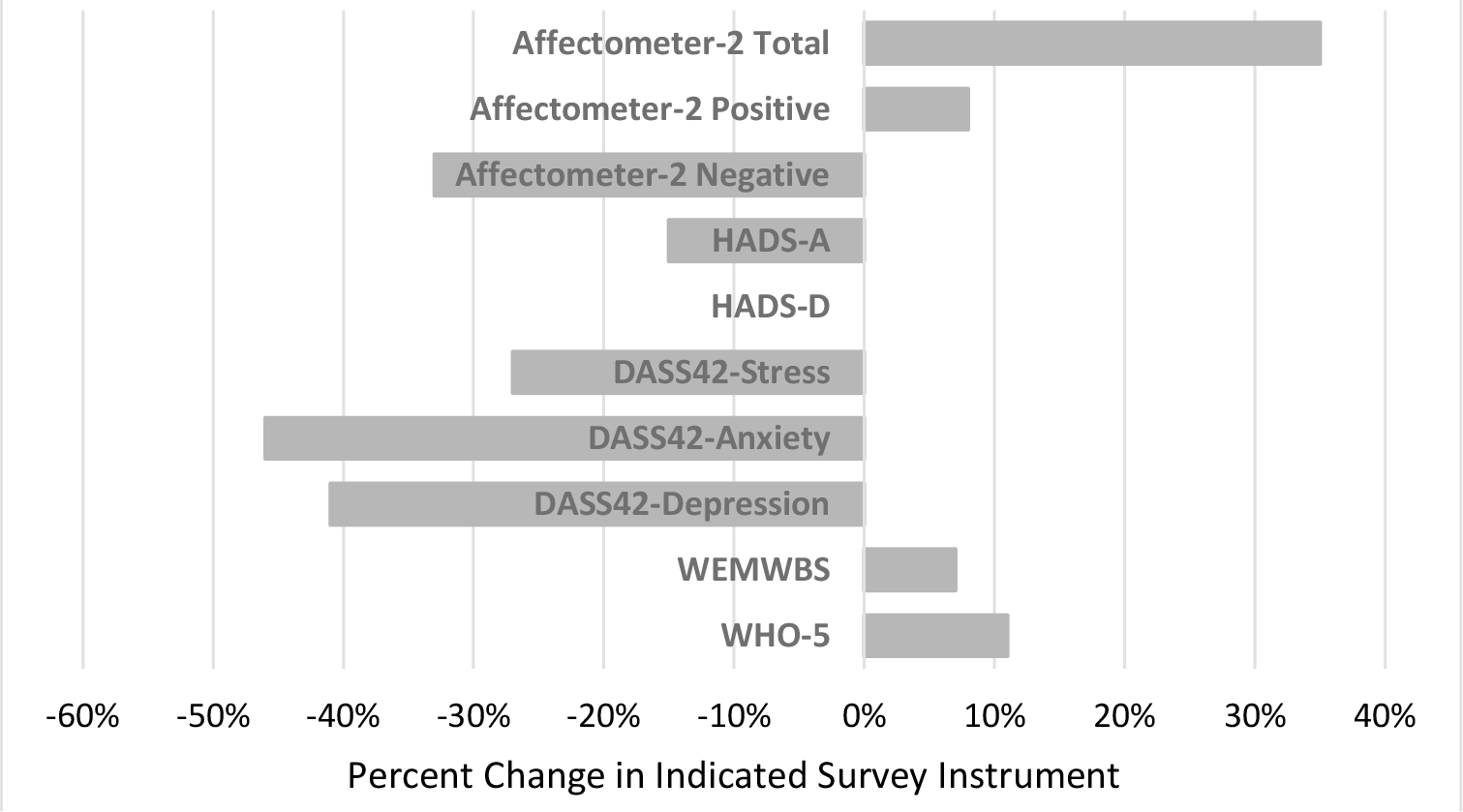
Changes in Mood and Happiness after Diet Change.

Unexpectedly during the course of the study, uBiome filed bankruptcy and stopped processing all microbiome samples. Prior to uBiome closing we obtained native sequencing files and proprietary uBiome diversity indices for each completed specimen (N=17). Due to uBiome’s shutdown, the study altered its original plan for gut diversity analysis by relying on Dr. Kraft and Dr. Rishishwar for analysis of the uBiome microbiome sequencing files. As previously planned, we calculated various standard microbial diversity indices in addition to the uBiome diversity index: Shannon, Simpson, Inverse Simpson, and Fisher diversity indices. In this group of completed microbiome diversity analyses shown in **Table 4**, none of the diversity indices showed a change from baseline to follow-up with significant p-values. However, we found that the uBiome gut microbiome diversity index varied significantly by gender, but not by race. The uBiome diversity index was higher in females than males (8.66 vs. 7.96, p=0.024) and remained significantly higher even after accounting for the two time points around the diet change (repeated measures ANOVA p=0.0748).

**Table 4.** Microbiome Characteristics of Enrolled Subjects.

| Table 4. Microbiome Characteristics of Enrolled Subjects |  |  |  |
| --- | --- | --- | --- |
| Characteristic | Baseline | Follow-Up | p-value |
| uBiome diversity index (mean, SD) | 8.5 (0.6) | 8.3 (0.7) | 0.499 |
| Shannon diversity index (mean, SD) | 3.9 (0.6) | 4.1 (0.4) | 0.616 |
| Simpson diversity index (mean, SD) | 0.94 (0.06) | 0.96 (0.02) | 0.467 |
| Inverse Simpson index (mean, SD) | 31.4 (17.8) | 32.8 (13.6) | 0.861 |
| Fisher diversity index (mean, SD) | 31.2 (13.5) | 29.4 (8.4) | 0.749 |

The final analysis included correlations between gut diversity, mood scores, and dietary composition. We first noted correlations between diet and mood, particularly for dietary proportions of fat and protein intake positively correlated with the happiness, wellness and well-being, and less anxiety and depression (see **Appendix Table 5** for the Pearson correlation coefficient [r] and p-value for each analysis). Specifically, fat and protein content of the diet negatively correlated with the DASS-42 Depression and Anxiety scores, and the HADS Anxiety scores (**Appendix Table 5**). While the analysis of mood and diet for all timepoints together found the strongest correlations between percent fat and percent protein, the analysis of the change in mood and the change in diet also found strong correlations between percent fat, percent carbohydrate, and percent fiber (**Appendix Table 6**). The change in HADS Anxiety scores with the change in percent fat illuminated a correlation of -0.6708 and a p-value of 0.0239, meaning this correlation has a mere 2.39% chance of being a total coincidence (**Fig. 4 & Appendix Table 6**). Furthermore, the HADS-Anxiety score also strongly correlated with the percent carbohydrates with a Pearson correlation coefficient r-value of 0.7859 (p-value of 0.0041), meaning changes in carbohydrate consumption account for 61.8% of the change we observed in the HADS-Anxiety score (r^2^=0.6176) (**Figure 5**).

**Figure 4.**
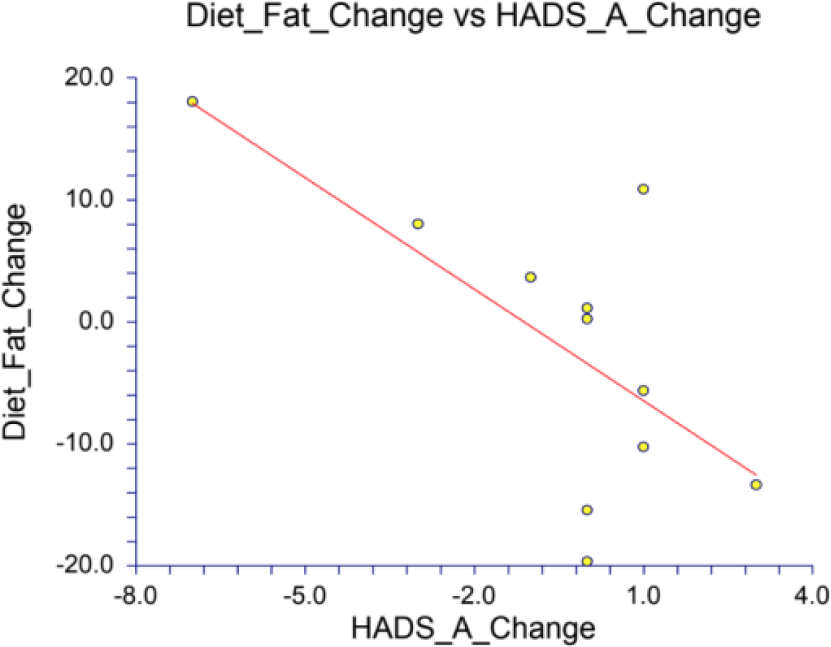
Change in HADS Anxiety correlation with change in percent fat (p-value of 0.0239)

**Figure 5.**
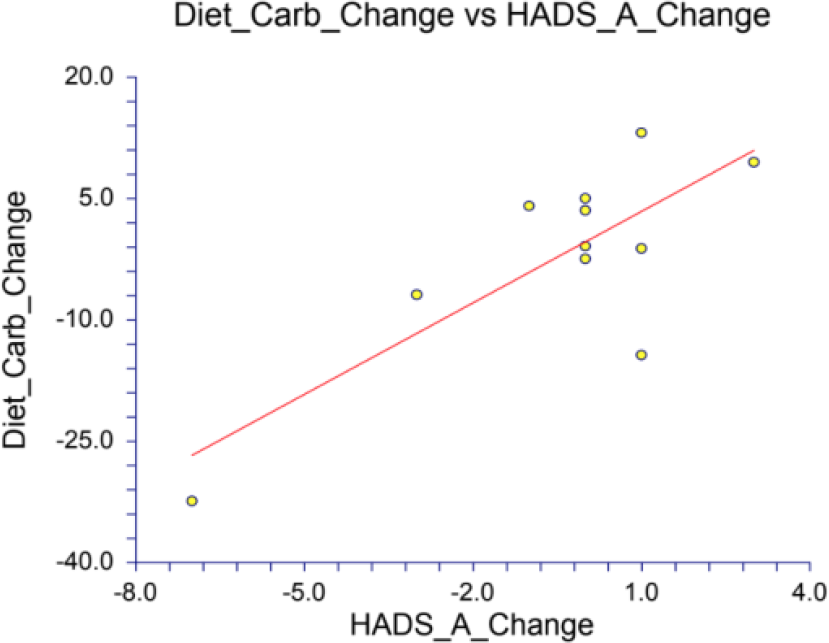
Change in HADS Anxiety correlation with change in percent carbohydrates (p-value of 0.0041)

The final analysis correlating diet with the gut microbiome also noted several important correlations. Correlations between dietary nutrient intake and the gut microbiome diversity were particularly apparent with total calories and total fiber: correlation between uBiome’s diversity index with total calories for both timepoints created a Pearson correlation coefficient r-value of -0.6619 (p-value of 0.0038) (**Appendix Table 7**). This was similarly observed with three other diversity indices: Shannon, Inverse Simpson, and Fisher produced p-values of 0.1549, 0.0966, and 0.0835, respectively. For fiber, the correlation with the uBiome diversity index was -0.4763 with a p-value of 0.0533, meaning a decrease in total fiber led to an increase in uBiome diversity (**Appendix Figure 6**). The correlation between diet and gut microbiome diversity remained strong when analyzing the changes in diversity with the changes in diet from the beginning to the end of the study. Specifically, the change in uBiome diversity score and the change in total calories yielded a Pearson correlation coefficient r-value of -0.6215 (p-value of 0.1000); and the change in uBiome diversity score and the change in total fiber intake yielded a r-value of -0.8002 (p-value of 0.0171) (see **Appendix Table 8**). All of the diversity indices were correlated with various dietary measures, and the correlations and p-values were placed in **Tables in the Appendix**.

Final analysis between mood and microbiome diversity showed strong correlations between particularly the HADS Anxiety and DASS-42 Depression and the various diversity indices. Results for the correlation of change in HADS-A with the change in the Shannon index display an r-value of -0.8111 and a p-value of 0.0501, meaning the gut microbial diversity increased while change in anxiety decreased (see **Appendix Table 9**). This means that the participants experienced less anxiety as their gut microbiome became more diverse. Furthermore, this remains true for the change in DASS-42 Depression correlation with the Shannon index, which produced an r-value of -0.7125 (p-value of 0.1121), meaning as the gut microbial diversity increased, the measures of depression went down (**Appendix Table 9**).

## Discussion

This pilot study examined the relationship between mood, microbiome diversity, and diet composition by analyzing 20 adults who underwent a substantial change in their diet. We found that diet changes that include higher percentages of fat and protein content led to increased well-being and decreased anxiety and depression while higher consumption of carbohydrates led to decreased happiness with greater anxiety and depression. We also found that decreased caloric and total fiber intake increased the gut microbiome’s diversity. Lastly, we discovered a correlation showing that as the gut microbiome becomes more diverse, anxiety and depression decrease.

Prior research on the gut microbiome has shown that diet affects both gut diversity and mood, typically within 24 hours. Studies on neurological diseases such as epilepsy have shown particular diets, like the ketogenic diet, improve the symptoms of the disease. This study furthers that earlier research by examining otherwise healthy individuals and showing the opposite side to those studies: diet’s effect on the mood and happiness rather than diet’s effect on mental and physical disorders. Consuming protein has been shown to reduce absorption of carbohydrates and release dopamine, while consuming carbohydrates releases serotonin, both of which directly affect the mood. This study furthers that information, displaying fat and protein’s positive effects on mood simultaneously with carbohydrates’ negative effect through anxiety and depression. Further research into this subject could determine the percentage of each dietary component that promotes the highest happiness and well-being, lowest anxiety, and lowest depression all together in one diet.

This study utilized several validated methods to characterize the microbiome and to assess mood and happiness, but as a pilot study this project was limited by a small number of study subjects. In addition, mood is subjective and varies from day-to-day, mood may be directly affected by diet without affecting the gut microbiome, and other factors may be involved which were not ascertained by this study. For mood, it is important to keep in mind the measures of each survey, as some measure positive characteristics while others track negative ones. The Affectometer-2 measures both positive and negative, and the total subtracts the negative from the positive; thus, a more positive score means a happier and more satisfied human being. The HADS measures anxiety and depression in two different scores, with a higher number meaning more anxious or more depressed. The DASS-42 measures stress, anxiety, and depression in three different scores, so a more positive number means more stress, anxiety, or depression. The WEMWBS is a measure of satisfaction and happiness, so a higher total score means a happier person. Finally, the WHO-5 is also a measure of happiness and satisfaction, so a higher percentage would be a happier person. Some limitations to this study beyond the subjective nature of the term “mood,” are compliance with following the study protocol, lack of exact control over the first and final diets, and uBiome’s bankruptcy that limited sample size further than expected.

This pilot study examined correlations between diet composition, mental health, mood and happiness, and the gut microbiome to find that fat and protein reduce anxiety and depression while carbohydrates have the opposite effect. Furthermore, total calories and fiber had a negative correlation with gut microbiome diversity, and anxiety and depression decrease as the gut diversity increases. Despite the limitations of the study, the results suggest that further research into the particulars of diet, mood, and microbiome could determine specific proportions of each dietary component to maximize satisfaction and minimize anxiety and depression in a personalized manner.

## Data Availability

All data produced in the present study are available upon reasonable request to the authors.

### Appendix

**Table 5.** Correlations between mood surveys and diet composition for all subjects & all timepoints.

| r/p-value | Total calories | % fat | % carb | % protein | Total fiber |
| --- | --- | --- | --- | --- | --- |
| Affectometer |  | 0.3385/0.0908 |  | 0.3431/0.0862 |  |
| Affectometer+ |  | 0.3547/0.0754 |  | 0.2995/0.1372 |  |
| Affectometer- |  | -0.3092/0.1243 | 0.2383/0.2411 | -0.3761/0.0583 |  |
| DASS-S |  |  |  | -0.2454/0.2269 |  |
| DASS-A |  | -0.3693/0.0634 |  | -0.3807/0.0550 |  |
| DASS-D |  | -0.4108/0.0371 | 0.2889/0.1523 | -0.4314/0.0278 |  |
| HADS-A |  | -0.4027/0.0414 | 0.2495/0.2191 | -0.3233/0.1072 |  |
| HADS-D |  |  |  |  |  |
| WEMWBS |  | 0.3006/0.1356 |  | 0.2505/0.2171 |  |
| WHO |  | 0.3587/0.0719 | -0.2681/0.1854 | 0.3381/0.0911 |  |

**Table 6.**
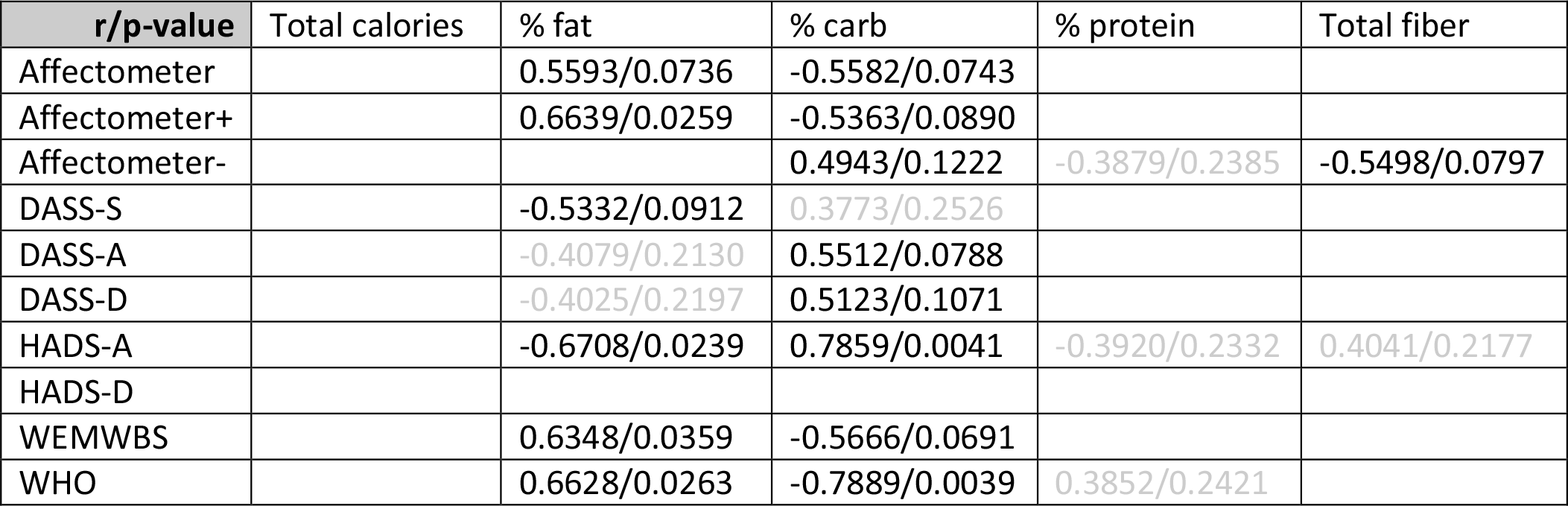
Correlations between mood changes and diet changes.

**Table 7.**
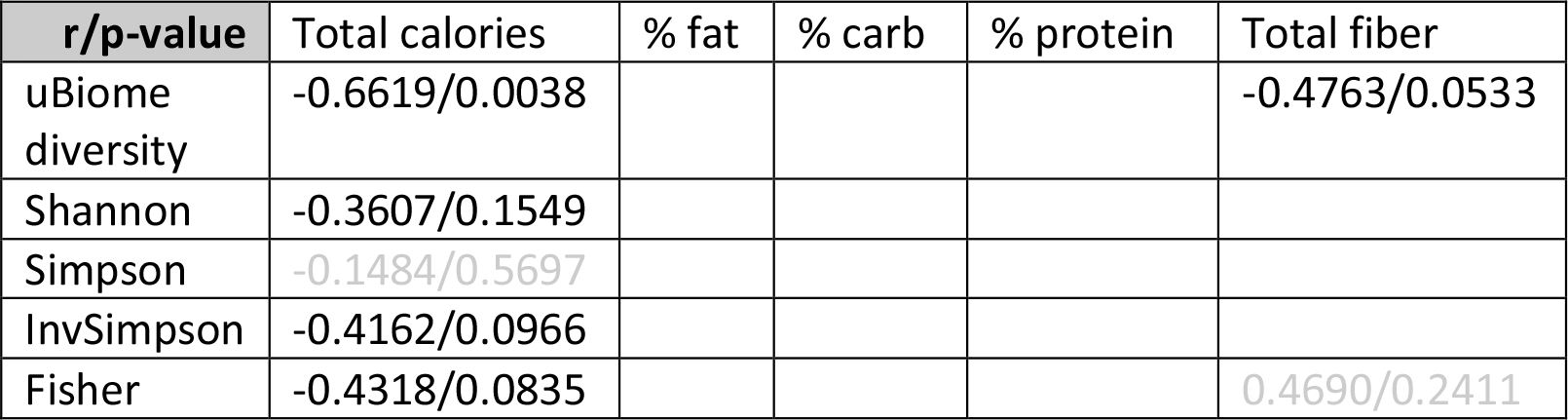
Correlation between diet and gut microbiome for all subjects & all timepoints.

**Figure 6.**
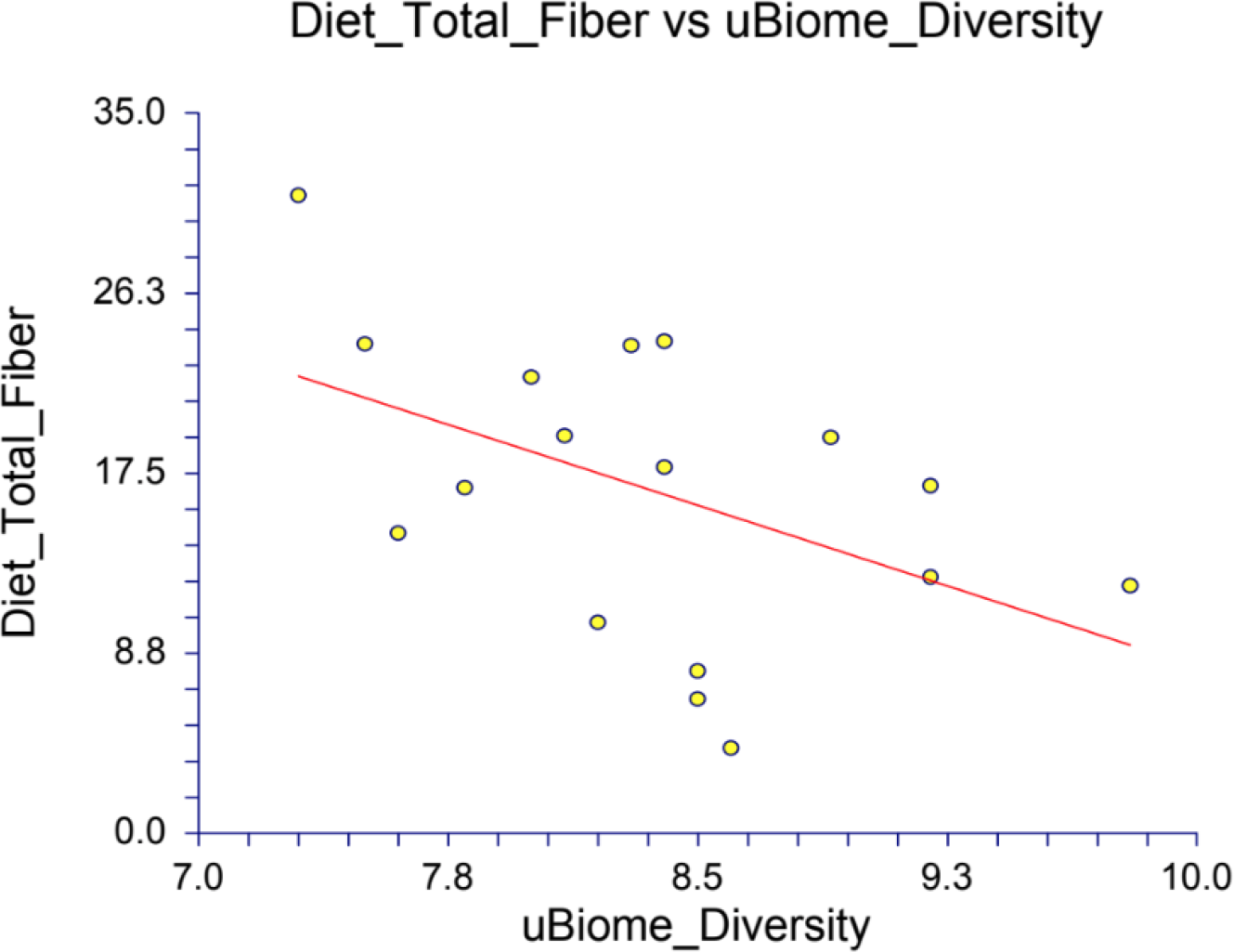
Correlation between uBiome diversity and total fiber for all timepoints (p=0.0533)

**Table 8.**
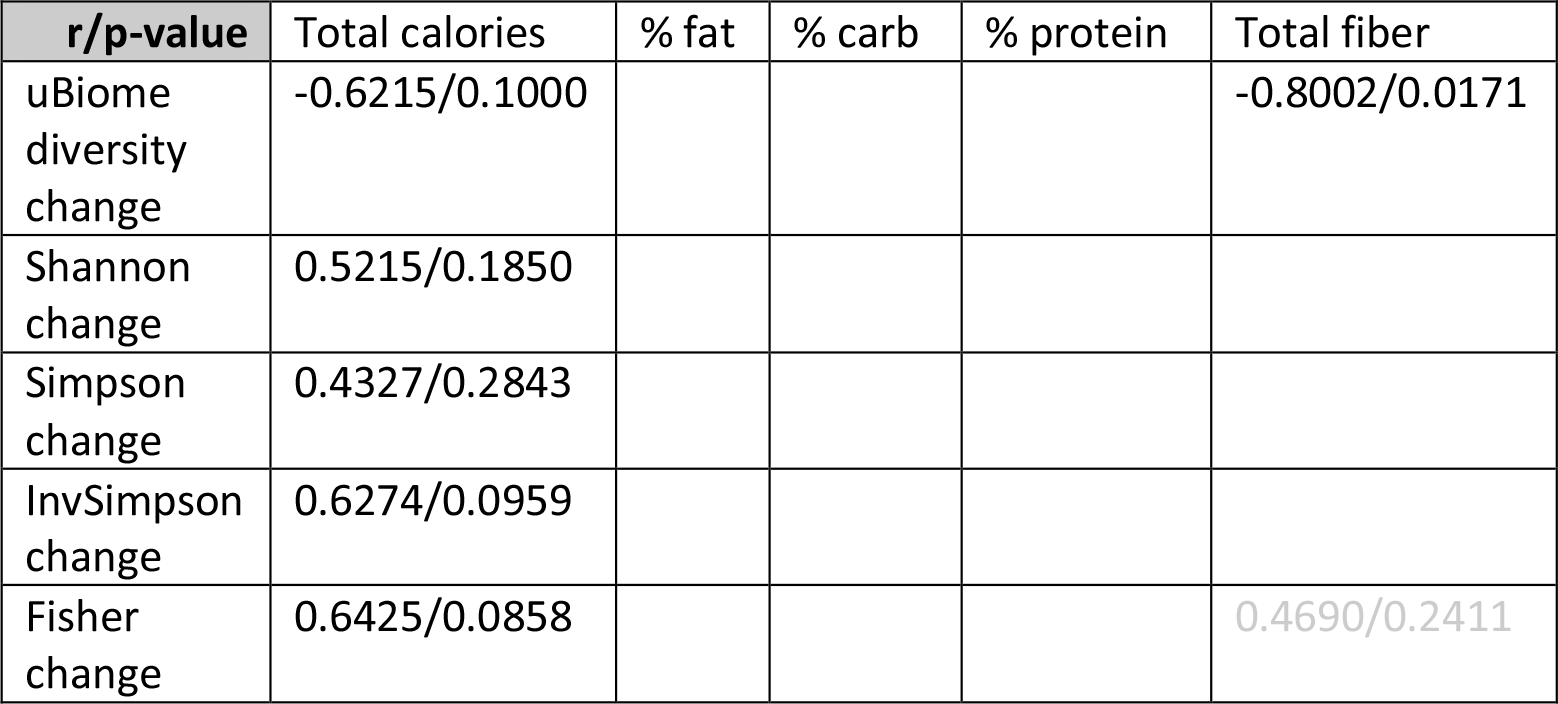
Correlation between changes in diet and changes in gut microbiome.

**Table 9.**
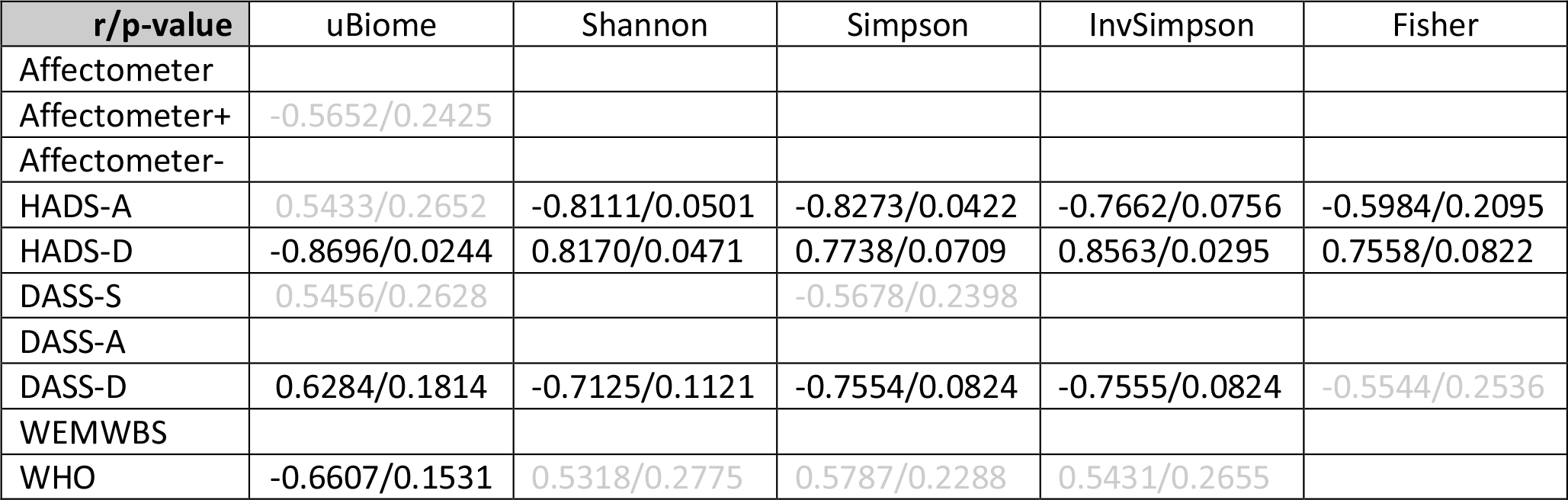
Correlations between changes in different diversity indices and the changes in mood surveys.

